# Underdiagnosed and undertreated peripheral arterial disease: Using design thinking to establish priorities for peripheral arterial disease care

**DOI:** 10.1101/2023.11.27.23298968

**Authors:** Radha Joseph, Sean Park, Teresa M. Chan, Vinai Bhagirath, Sonia S. Anand

**Affiliations:** Department of Medicine, Faculty of Health Sciences, McMaster University; Office of Continuing Professional Development, Faculty of Health Sciences, McMaster University; Population Health Research Institute

## Abstract

**Introduction:** Design thinking (DT), a methodology for solving complex problems, has the potential to create powerful, human-centred healthcare improvement. We applied DT methodology to the context of peripheral arterial disease (PAD). PAD is increasingly prevalent globally and associated with significant morbidity and mortality. We fall short of achieving effective secondary prevention due to persistent underdiagnosis and undertreatment of this disease. In this study, we sought to identify novel and creative solutions to improve diagnosis and secondary prevention of PAD.

**Methods:** We describe the initial ‘Empathize’, ‘Define’, and ‘Ideate’ stages of the five-stage DT model proposed by the Hasso Plattner Institute of Design at Stanford University. We engaged patients with PAD, caregivers, clinicians, and other stakeholders in a co-design process using semi-structured interviews, a DT workshop, and post-workshop survey. Data from the interviews and workshop were analyzed using inductive thematic analysis, and data from the survey were analyzed using an idea prioritization matrix.

**Results:** Exploring the lived experience of those with PAD and those delivering PAD care emphasized the influence of system-level barriers. Many of the solutions proposed by workshop participants target evidence-based, system-level interventions through improved funding support, institutional support, outreach efforts and technological applications. The connections between insights derived in the ‘Empathize’ stage and solutions proposed during the ‘Ideate’ stage showed the success of the co-design process in inspiring empathy-driven solutions.

**Discussion:** This study demonstrates how DT methodology can be applied to complex healthcare problems such as PAD care, to systematically develop human-centred solutions. In the next stages of this study, we will use the results of this co-design process to iteratively implement, evaluate, and optimize the proposed solutions which were prioritized as being most feasible and high impact.

**KEY MESSAGES:** *What is already known on this topic:* Peripheral arterial disease (PAD) is increasingly prevalent globally. The significant morbidity and mortality associated with PAD can be reduced with timely diagnosis and the effective use of secondary preventative therapies; however, PAD remains underdiagnosed and undertreated compared to other atherosclerotic diseases.

*What this study adds:* This study is novel in its application of design thinking methodology and a co-design approach to work together with people with lived experience of PAD, to establish priorities for PAD care. How this study may affect research, practice or policy – Insights from this study emphasize system-level barriers which prevent effective delivery and uptake of PAD care. Solutions that are human-centred and co-produced with patients and key stakeholders should improve institutional and governmental support for implementation of evidence-based best practices; this will be investigated further in the next stages of this study.

## INTRODUCTION

The public healthcare system in Canada is in a state of disarray, a challenging state due to a “complex set of problems with inextricable interdependencies.”(1) Many of these problems could be characterized as “wicked problems”, complex problems that defy analytical problem solving, are not understandable by any single individual, and have no single best solution.(2,3) A traditional top-down approach to problem-solving involves data evaluation, proposal of a solution, and deployment of this solution to users; however, this offers a very narrow window into ambiguous problems. Furthermore, if there is insufficient collaboration with stakeholders, especially patients and providers, then the deployed products or services may remain unused, because they do not account for human context, need, or imperfection.(4) This is especially true with regards to underserved populations, whose unique needs are often overlooked. To bridge this gap between intervention development and implementation, we must be human-centred and creative in our improvement and innovation efforts. As such, Design Thinking (DT) methodology is a powerful tool for tackling wicked problems.(5,6)

In this study, we used DT to investigate the wicked problem that is the persistent underdiagnosis and undertreatment of peripheral arterial disease (PAD).

### The context of our problem

PAD is an increasingly prevalent global problem that is estimated to affect more than 230 million people worldwide.(7,8) Lower extremity PAD is an atherosclerotic disease of the arteries supplying the legs, which causes symptoms ranging from intermittent claudication (leg pain with exertion) to critical limb ischemia (or chronic limb threatening ischemia, characterized by resting leg pain and tissue loss).(9,10) Across the spectrum of disease, PAD is associated with significant clinical, functional and psychological consequences.(11) Patients with PAD are at risk of major adverse cardiovascular events, major adverse limb events and mortality.(12,13) In Canada, one year after revascularization for critical limb ischemia, more than 35% of patients undergo amputation or die.(14)

The morbidity and mortality associated with PAD can be reduced with timely diagnosis and the effective use of secondary preventative therapies including lifestyle modification, especially smoking cessation, and medications such as blood thinners, cholesterol-lowering drugs, and blood pressure-lowering drugs.

Unfortunately, PAD is underrecognized compared with other atherosclerotic diseases; this lack of awareness has contributed to underdiagnosis of this disease.(15) Furthermore, PAD is undertreated. Compared to patients with coronary artery disease, patients with PAD derive equal or greater benefit with secondary preventative therapies; however, around the world, patients with PAD are significantly less likely to use or be prescribed these therapies. This may be due to implementation barriers occurring at the level of the patient, health care provider, or health system; examples include poor awareness and understanding of PAD as a diagnosis and disease, cost of medications, lack of clarity regarding which healthcare provider is responsible for optimizing secondary prevention, and lack of access to comprehensive secondary prevention programs.(16) Barriers to appropriate, effective and timely care are particularly consequential for patients with low socioeconomic status and/or residing in rural areas, who have higher rates of presentations with complications, higher rates of emergency surgeries, and higher amputation rates.(17,18)

### Rationale

Ultimately, these issues highlight the need to transform how PAD care is designed and delivered. The traditional process of designing *for* or *at* people–where researchers, healthcare providers, and administrators design new models of care based on research studies and expert opinion–has not led to improvement in the underdiagnosis and undertreatment of PAD. Therefore, DT methodology and a co-design approach was chosen as a means of disrupting the traditional processes and designing *with* people–allowing the perspectives of patients and key stakeholders to be integrated into key stages of the process.

In this article, we describe our use of co-design and DT methodology to identify novel and creative solutions to improve diagnosis and secondary prevention of PAD in our local, provincial, and national context. By bringing together patients, caregivers, healthcare providers and healthcare leaders, we sought to collectively identify important insights and opportunities that could guide the development of desirable and feasible prototypes for implementation.

This article will discuss our findings from the ‘Empathize’, ‘Define’, and ‘Ideate’ stages of the DT process. The ‘Prototype’ and ‘Test’ stages will be further described and evaluated in a future article.

## METHODS

### Research approach

Healthcare is increasingly applying design knowledge and competence to deal with challenges(22).

DT has been defined as “a human-centred approach for solving complex problems employing attributes such as creativity, user involvement, multidisciplinary teamwork, iteration, prototyping and user centredness”.(23) We selected DT methodology because it prioritizes deep empathy for explicit, tacit and latent end-user desires, needs and challenges to understand how a problem is experienced in the context of people’s lives; this serves as the foundation for developing insights about where the opportunities and barriers to change are and potentially comprehensive and effective solutions which can be rapidly prototyped and tested.(20)

We engaged in co-design as a research philosophy and practice that aims to empower researchers and participants, service users and service providers, and other stakeholders to share power in various aspects of the design process; this includes making decisions about important themes, generating ideas, and making sense of data.(19)

This process moves through stages of analysis, synthesis, iterative prototyping and testing, to implementation.(20) The process is often taught as a stepwise, staged methodology; however, the various activities and methods can be used in a variety of combinations depending on the context and needs of the project. In practice, the process is often non-linear as involving patients in generating or testing out new ideas yields deeper insights about their needs and experiences, which can then shift what is considered a challenge or opportunity. (24)

We followed the five-stage DT model proposed by the Hasso Plattner Institute of Design at Stanford University (the d.school): ‘Empathize’, ‘Define’, ‘Ideate’, ‘Prototype’, ‘Test’ (**Figure 1**).(21)

**Figure 1.**
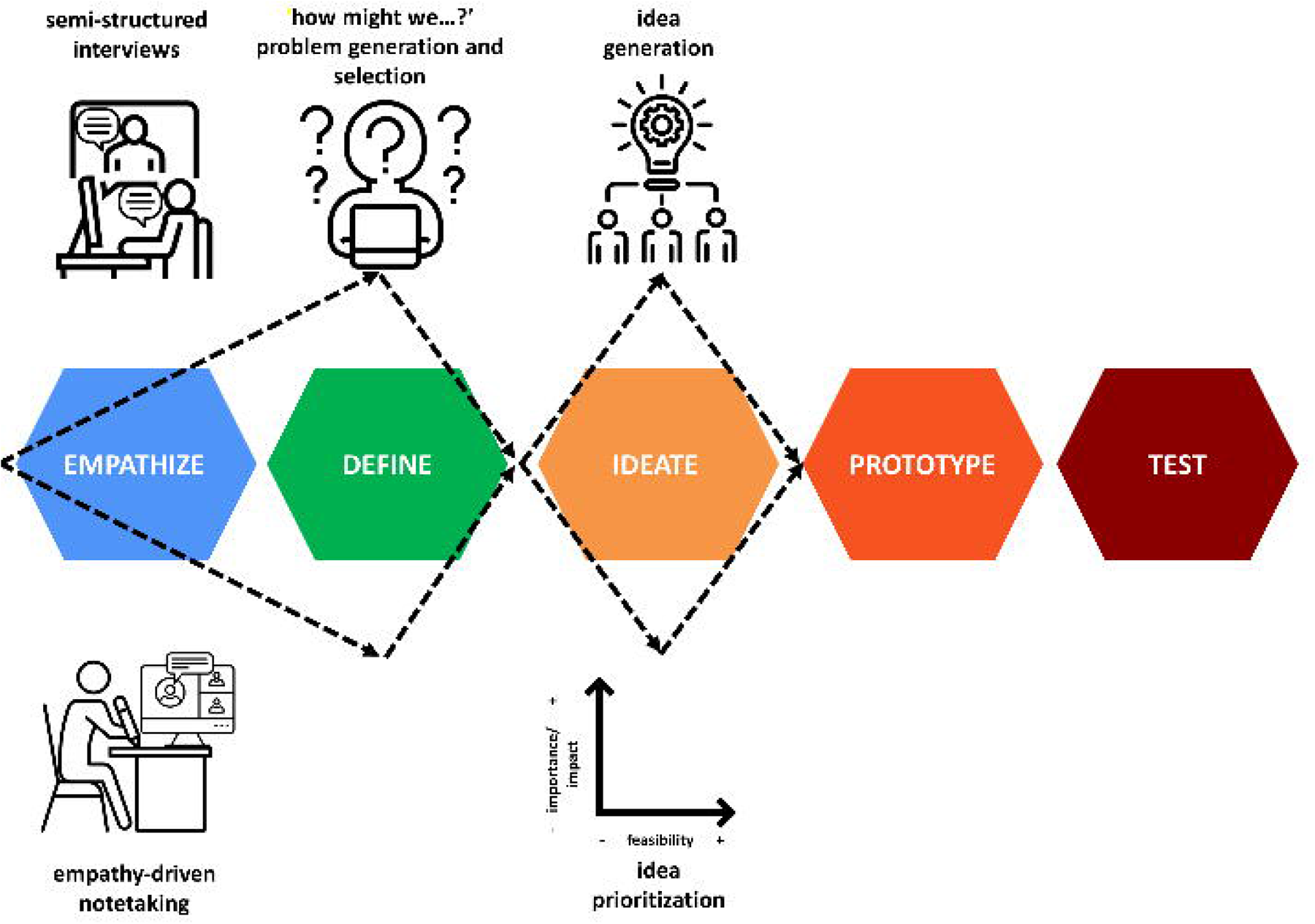
The five-stage design thinking (DT) model proposed by the Hasso Plattner Institute of Design at Stanford University (the d.school), adapted to illustrate our research process and the various phases of divergence and convergence. In the ‘Empathize’ stage, the needs and challenges of patients with peripheral arterial disease, their caregivers, and their healthcare providers were explored using a) semi-structured interviews with patients, caregiver and clinicians, and b) empathy-driven note taking during a DT workshop. In the ‘Define’ stage, insights gathered during the ‘Empathize’ stage were the foundation for ‘How might we…?’ problem generation and selection. In the ‘Ideate’ stage, participants brainstormed creative solutions to the selected ‘How might we…?’ problem, which were later prioritized by feasibility and importance/impact using an idea prioritization matrix. The ‘Prototype’ and ‘Test’ stages will be further described in a future article.

### Study setting

This study was performed in 2022 at McMaster University, in Hamilton, Ontario, Canada, a tertiary care center and network of teaching hospitals. Care for people with PAD at McMaster includes diagnostic imaging, surgical and endovascular therapies, an inpatient ward, and outpatient clinics. Vascular Surgery, Vascular Medicine, and Thrombosis physicians collaborate to provide care for these patients at McMaster.

### Ethical considerations

As a needs assessment for a quality improvement process, an exemption was granted by the Hamilton Integrated Research Ethics Board after a review of the intended process. As per national policy for such projects, written consent from participants was not required; however, participants were aware of the researchers’ intention to be scholarly in the dissemination of findings from this project.

### Research team characteristics and reflexivity

The research team included two project leaders, one Vascular Medicine trainee, and two design consultants from McMaster University. One project leader is a Vascular Medicine clinician and researcher, with a focused area of interest in vascular diseases, clinical trials, and population health studies; the other is a Thrombosis clinician and researcher, with a focused area of interest in arterial diseases. The trainee was in the Vascular Medicine fellowship program at the time the workshop was conducted. One design consultant is an emergency physician, clinician educator, and education scientist; the other is a health design educator and researcher.

### Participant selection and engagement

The research process involved semi-structured interviews and a ‘Design Thinking Workshop’, as detailed below.

The semi-structured interviews were conducted by a design consultant with 3 patient partners, 1 caregiver and 2 clinicians.

The 3 patient partners were selected based on their disease severity and presence/absence of caregiver supports. All three patients identified as male; a female patient was also approached, but declined to participate as she felt that she was always busy attending appointments and in and out of hospital because of PAD, which she described as a burden. All 3 patients had a diagnosis of PAD, and 2 patients had undergone lower extremity amputation as a result of PAD. For 1 patient, his spouse/caregiver was highly involved in supporting him, therefore she was invited to participate.

The 2 clinicians were selected given their expertise and clinical practices caring for patients with PAD.

The ‘Design Thinking Workshop’ was attended by all individuals who participated in the semi-structured interviews. Invitations were also sent to key stakeholders, including patients, family members, clinicians (surgeons, physicians, nurses, and pharmacist), researchers, and healthcare leaders and administrators. A total of 21 people attended, including Vascular Surgeons and a Vascular Surgery trainee, Vascular Medicine specialists and trainees, an Internal Medicine specialist, nurses, pharmacists, postdoctoral fellows, and a hospital leader.

### Process and timeline

Here, we describe three stages of our design-thinking process: ‘Empathize’, ‘Define’ and ‘Ideate’.

#### I. Empathize

Empathy is at the core of human-centered design processes including DT.(21) In order to design solutions for patients with PAD, we explored the needs and challenges of patients with PAD, their caregivers and their healthcare providers.

For the ‘Empathize’ Stage, we engaged 3 patient partners, 1 caregiver and 2 clinicians in semi-structured interviews. Due to the COVID-19 pandemic, the interviews were conducted virtually using Zoom, a cloud-based video conferencing service. With the goal of eliciting their lived experience of PAD across their care journey, rather than a single point of care, patients and caregiver were asked to narrate their experience of living with PAD, and to share the biggest challenges and concerns they face in their daily lives. They were also asked to reflect on change they desire in the healthcare system. Clinicians were asked to discuss ‘pain points’ (challenges and barriers) that they encounter when providing care to patients with PAD.

Interview data were qualitatively analyzed using inductive thematic analysis.(25,26) Ideas and quotes from the interviews were anonymized and aggregated by theme. The research team reviewed the themes and developed insight statements. An insight statement is a short sentence that represents user perspectives, motivations, and tensions from the thematic data to define a human need.(27) This approach is specific to human-centered design, and novel to the academic literature for qualitative data analysis.(28) The goal of developing insight statements is to ascribe meaning to thematic data.(29)

A half-day, 3-hour ‘Design Thinking Workshop’ was then held. Again, owing to the COVID-19 pandemic, this was conducted virtually using Zoom. Workshop participants were provided with an overview of the DT process. Participants then engaged in some empathy work with each other. Insight statements from the semi-structured interviews were shared with participants. They then observed conversations between the workshop organizers, patients and caregiver, and clinicians. Participants were encouraged to use empathy-driven notetaking to capture their thoughts on what the interviewees were saying, thinking, and feeling during these conversations. Participants were encouraged to use an online word processor, Google Docs, for notetaking, to share their findings with the other participants and capture these thoughts in a visual, shareable format to refer back to over the course of the workshop.

#### II. Define

The goal of the ‘Define’ stage is to delineate the challenge that needs addressing. This is done with an understanding of the person or people you are designing for, needs that are important to fulfill, and insights gathered through empathy work.(21) Workshop participants were assigned to 3 groups, and asked to craft ‘How Might We’ (‘HMW’) questions. ‘HMW’ questions frame specific opportunity spaces for addressing identified needs and open up the imagination for thinking later on about ideas and actions. ‘HMW’ questions were recorded using Google Docs. Through this exercise, participants reflected on the insights gained through the ‘Empathize’ stage, and reframed the challenges experienced by patients, caregivers, and clinicians into opportunities to innovate.

Each group was asked to select the single most important HMW question to share with the larger group. All 3 groups then converged and agreed upon the most important HMW question, thereby defining and making explicit a meaningful and actionable problem statement.

#### III. Ideate

The ‘Ideate’ stage is the process of generating solutions to address the challenge identified during the ‘Define’ stage.(21) Initially, the goal is to push for the highest quantity and widest breadth of ideas, not simply finding a single, best solution.(21)

Workshop participants were assigned to 3 groups, given guidelines for how to brainstorm effectively, and asked to brainstorm solutions to the selected ‘HMW’ question. Solutions were recorded using Google Docs. Participants were encouraged to think outside the constraints of the existing problem space; for example, they were asked to consider what they would propose if cost was not an issue. They were also asked to defer judgment, or separate the generation of ideas from the evaluation of ideas; this is intended to fuel creativity, understanding that merits of each idea are assessed later.(21)

The solutions from all 3 groups were aggregated. The research team then refined, analyzed, and categorized solutions by theme using a general inductive approach. This refined list of solutions was used to populate a survey administered to all workshop participants. Participants were asked to vote about each idea along 2 axes: 1) feasibility–how much effort they perceived this initiative would take; 2) importance/impact–how much urgency and need they perceived for the proposed initiative, and how much value they perceived it would add to the PAD community. Based on the survey results, a prioritization matrix was created to display feasibility (effort) vs. priority/importance/impact.

This prioritization matrix will guide selection of ideas for the next stages, ‘Prototype’ and ‘Test’. The ‘Prototype’ and ‘Test’ stages will be further described and evaluated in a future article.

### Patient and public engagement

Patients and a caregiver were first involved in the data collection phase of this research, specifically through their participation in semi-structured interviews, the DT workshop, and post-workshop survey. As detailed above, the use of DT methodology ensured that their experience, priorities and preferences informed the research question/problem definition, and prioritization of future innovation ideas. They were provided with a stipend for their participation. These patient and public partners will continue to be invited to participate in the next stages of this project, as we prototype and test solutions that emerged from the DT workshop.

## RESULTS

The insights, challenges and solutions that emerged from the first three stages of our design-thinking process, ‘Empathize’, ‘Define’ and ‘Ideate’, are described.

### I. Empathize

To better understand the experience of living with PAD, we explored the needs and challenges of patients and a caregiver through semi-structured interviews and observed interviews with empathy-driven notetaking. Insights that emerged from this process fall broadly under 6 themes: lack of awareness about PAD in primary care; sense of self-infliction; functional and psychosocial consequences of PAD; mindset and motivation; family, social, and mental health supports; and openness, connectedness, and timeliness of interactions with the healthcare system (**Table 1**).

**Table 1.**
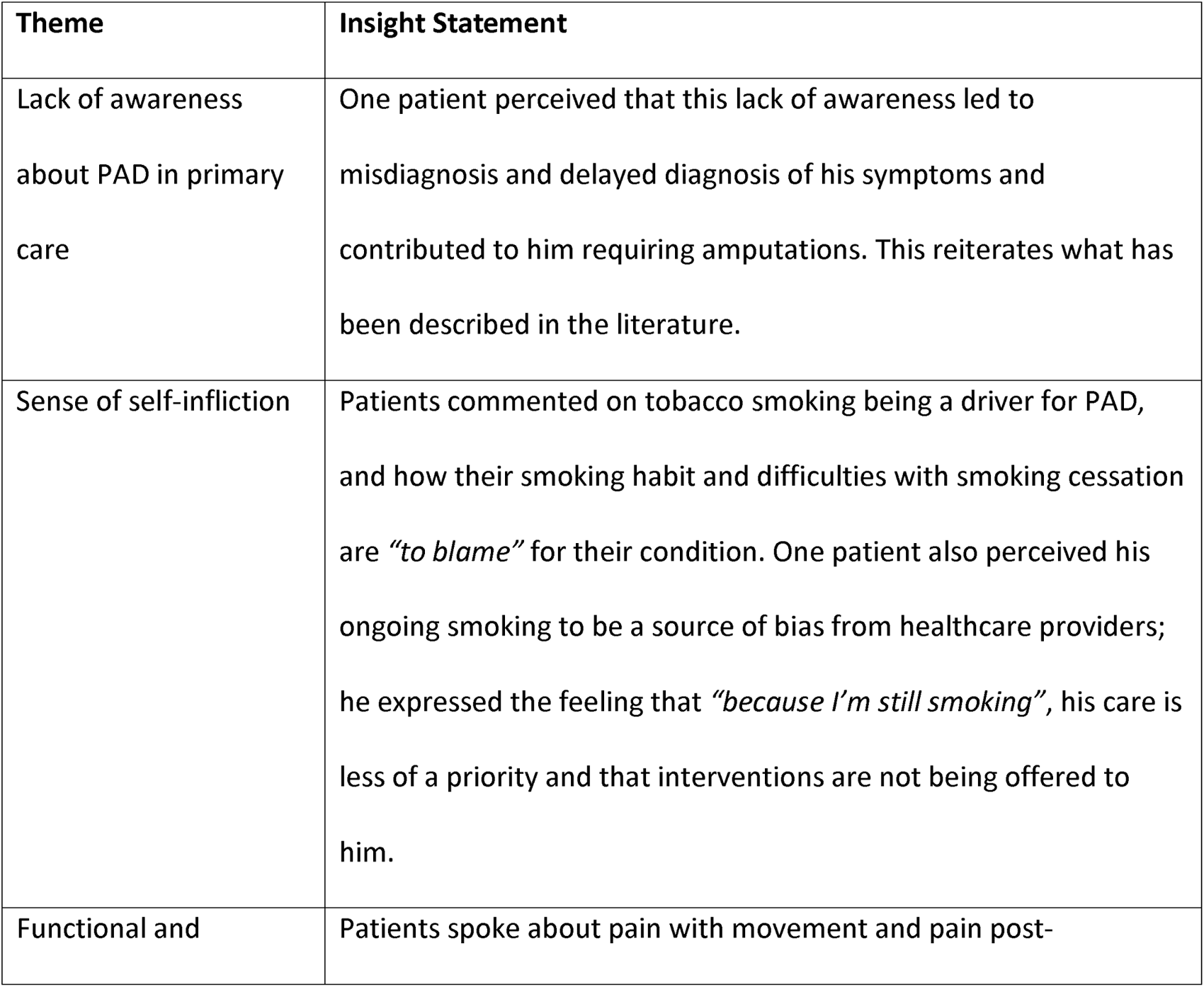
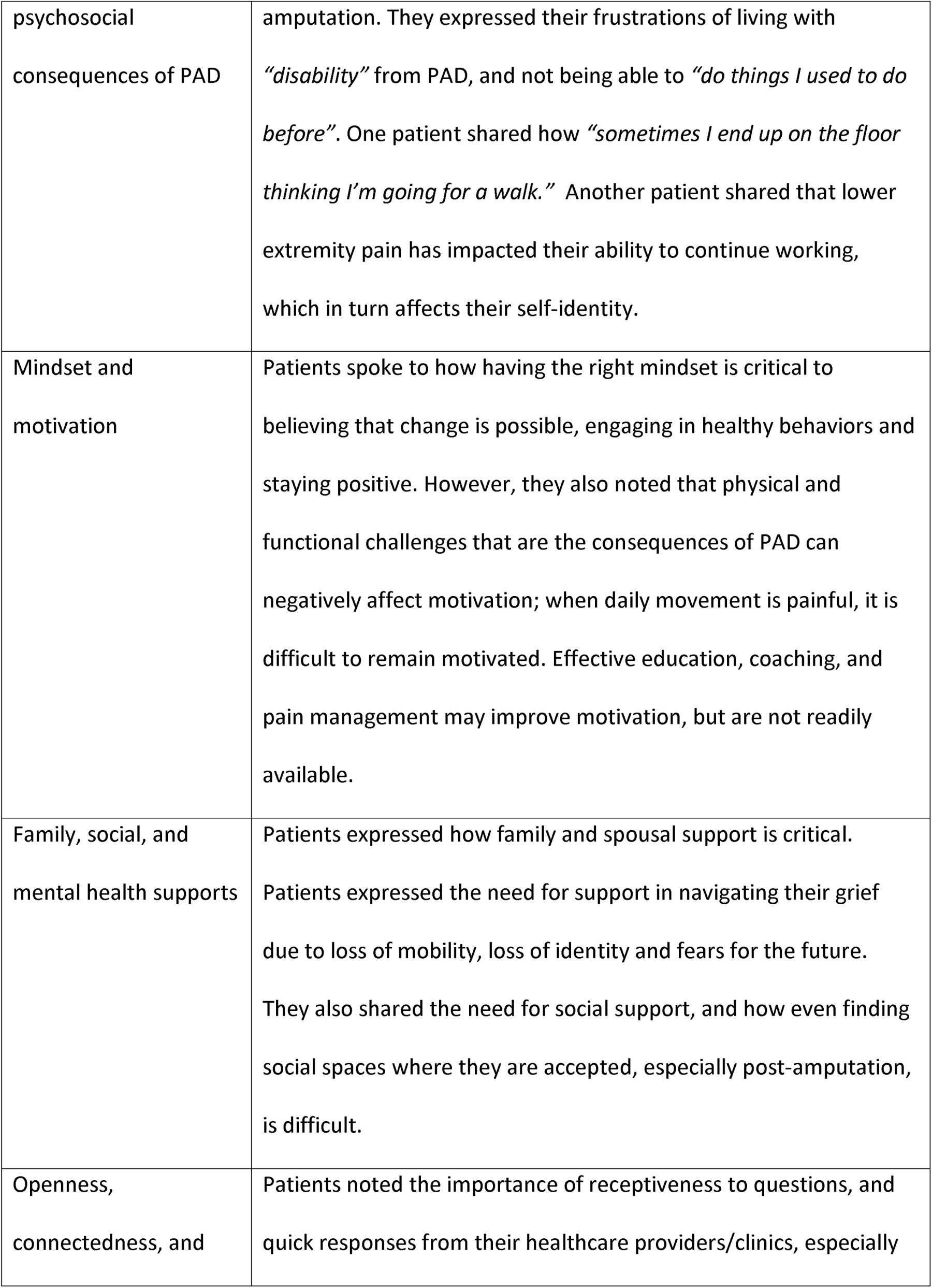
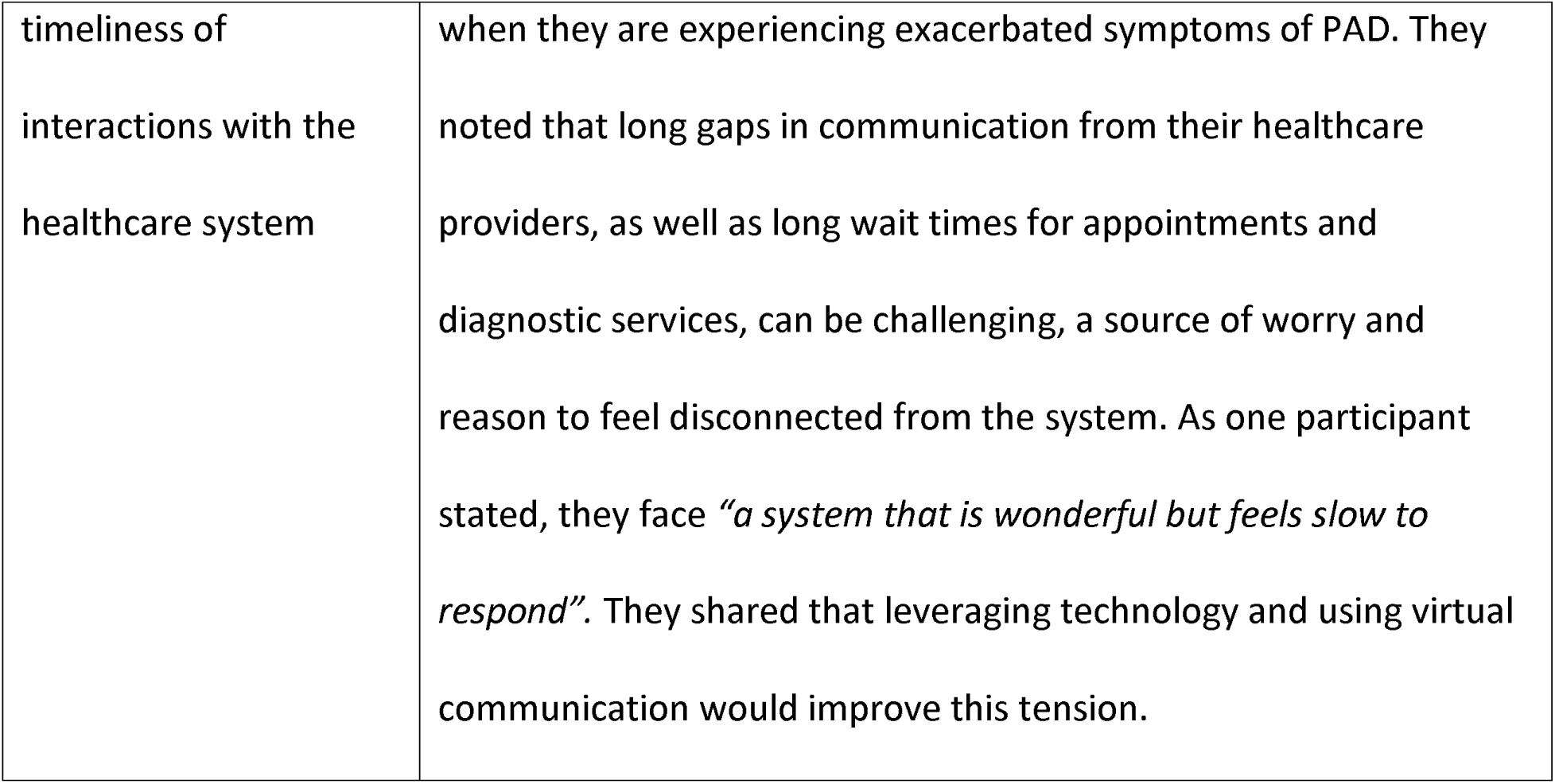

We also explored challenges for clinicians, again through semi-structured interviews and observed interviews with empathy-driven notetaking. Insights that emerged from this process fall broadly under 4 themes: time pressures; empathy for patients having to navigate the healthcare system; limited education, resources, and system support; and continued patient advocacy (**Table 2**).

**Table 2.**

| Theme | Insight Statement |
| --- | --- |
| Time pressures | Clinicians noted that they have very limited time to spend with patients due to high patient volumes (between 15-60 patients per day depending on specialty). They commented on the tension of wanting to spend more time with each patient versus the *“pressure of needing to see the next patient”*. |
| Empathy for patients having to navigate the healthcare system | Clinicians feel that the current healthcare system is *“not designed for patients”*. They see it as being *“patchwork”* and difficult for patients to navigate, with too much of the onus placed on patients (i.e., to navigate appointments for clinic visits, tests, etc.). |
| Limited education, resources, and system support | Clinicians expressed their disappointment with the healthcare system, with the under recognition of PAD compared to other atherosclerotic diseases; there continues to be relatively little education on PAD during medical training, and there are no resources to support preventative care programs. They expressed a sense of guilt that patients with PAD have been *“let down”*, and *“moral injury”* at knowing how to treat these patients but not always being able to provide that care. |
| Continued patient advocacy | Clinicians spoke of how they are *“not trained to change the system”*, and do not always feel welcome in the decision-making process. However, they see the need to *“band together”* and continue advocating for their patients, understanding that they *“can’t just wish for something to happen”*. |

### II. Define

‘HMW’ questions were proposed to delineate the challenge that needs addressing. A total of 36 HMW questions were brainstormed between the 3 groups, grounded in the insights gathered through empathy work. Each group was asked to select the single most important HMW question to share with the larger group, and then all 3 groups then converged and agreed upon the most important HMW question: *“How might we help patients to have early, easy access to health care advice and treatment, because we want to address the disease throughout the spectrum, not just at the urgent/late stage?”*

Therefore, the meaningful and actionable challenge was defined as improving timely access and ease of access to PAD care and treatment.

### III. Ideate

Workshop participants brainstormed solutions to the ‘HMW’ question above. The research team then aggregated, refined, analyzed, and categorized these solutions. After collapsing or combining similar ideas, 23 unique solutions remained, in 6 broad categories: technology-enhanced opportunities; streamlining patient experience; political changes; outreach; clinician education; and PAD patient support (**Table 3**).

**Table 3.**

| **Solution Category** | **Description** |
| --- | --- |
| Technology-enhanced opportunities | These solutions propose ways in which technology can be leveraged to develop or expand applications which will promote PAD awareness, education, behavioral change, etc., and improve access to existing services. |
| Streamlining patient experience | These solutions are aimed at simplifying and consolidating care for patients and referring healthcare providers, by way of a single point |
|  | of contact for systems navigation, a centralized, accessible, multidisciplinary care center, etc. |
| Political changes | These solutions look to increase recognition and prioritization of PAD and preventative care services—for example, through national and provincial lobbying. |
| Outreach | These solutions include education campaigns to increase PAD awareness amongst the public, and programs that would expand the reach of diagnostic and treatment services (i.e., pharmacy-based screening programs, mobile clinics, etc.). |
| Clinician education | These solutions are aimed at increasing PAD awareness amongst healthcare providers, from medical students to practicing healthcare professionals. |
| PAD patient support | This solution looks to expand supports for patients with PAD, by creating a space for patients at different stages in the PAD disease process to share their stories and experiences, facilitated by people with lived experience and/or healthcare professionals. |

Workshop participants were then sent a survey and asked to vote about each of the 23 solutions, along two axes: i) feasibility, or effort required to implement the idea; and ii) desirability, or importance and value of this idea for the PAD community. Thirteen of 21 (62%) participants responded to the prioritization survey, including 1 of the 3 patient partners. Most responses were from clinicians, researchers, and learners, plus 1 respondent from each category of teacher/educator, administrator/leader, research program manager.

Of the 23 solutions, the ideas which ranked highest with regards to feasibility and desirability on the prioritization matrix (**Figure 2**) are: PAD referral tool; “PAD hotline”; PAD systems navigator; PAD patient support group; and medical student education (**Table 4**).

**Table 4.**
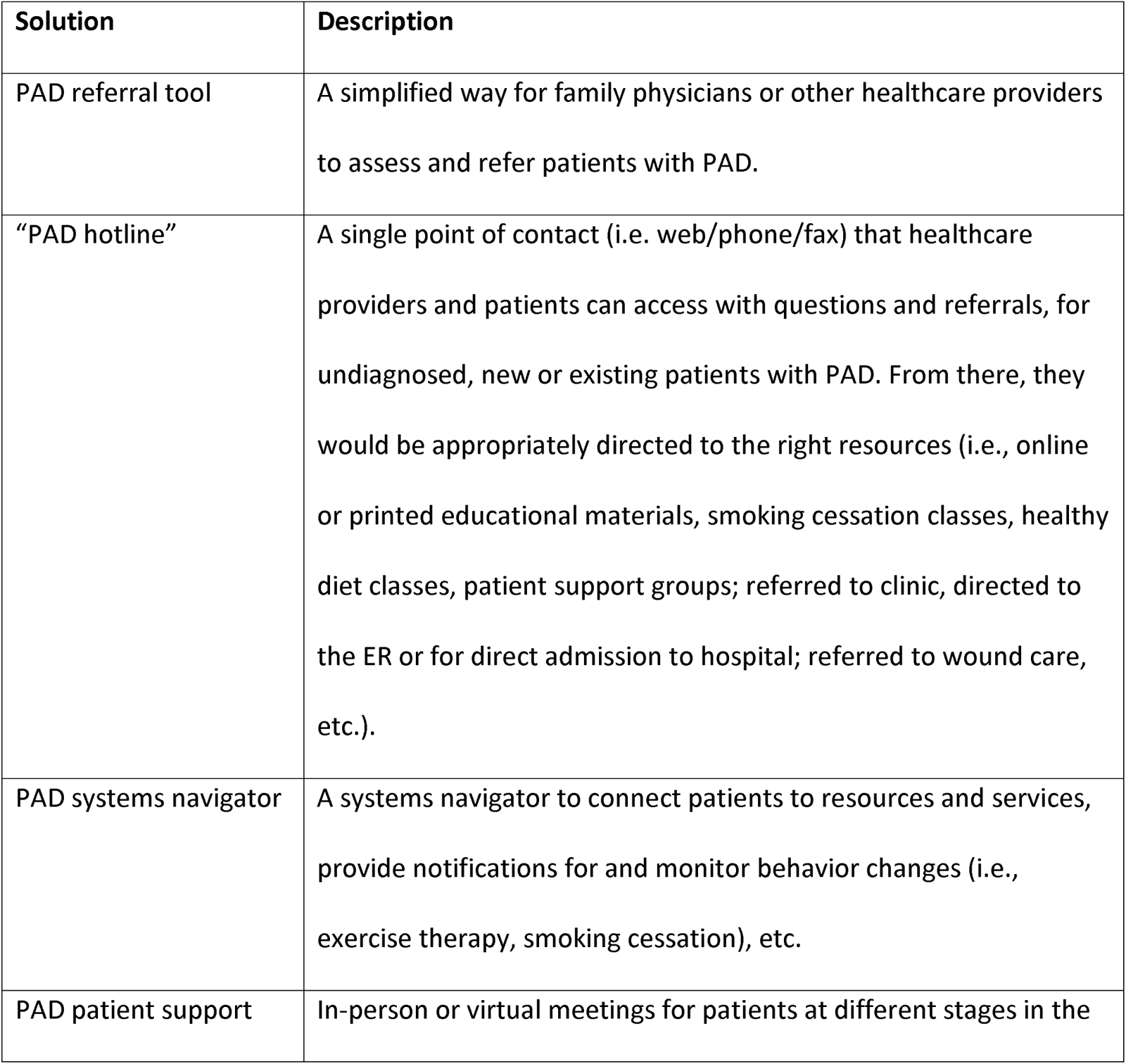
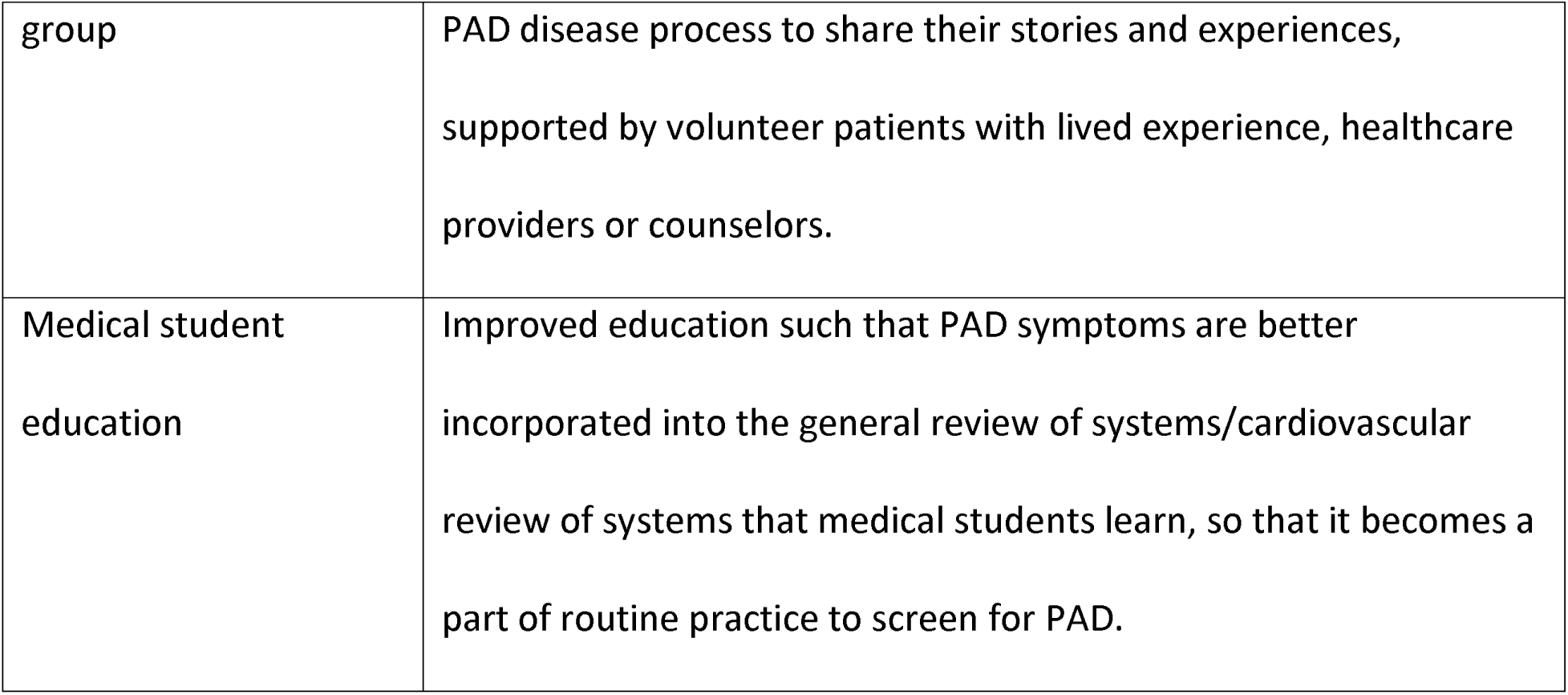

The prioritization matrix will guide selection of ideas for the next stages, ‘Prototype’ and ‘Test’.

## DISCUSSION

Globally, PAD is an increasingly prevalent condition, one with potentially severe consequences. Despite this, PAD remains underdiagnosed and undertreated. Our aim was to use DT methodology to identify novel and creative solutions to improve diagnosis and secondary prevention of PAD in our local, provincial, and national context.

Existing literature on challenges in PAD care acknowledges implementation barriers at the level of the patient, health care provider and health system.(16) In exploring the needs and challenges of those with lived experience of PAD, and clinicians involved in their care, there is an overwhelming preponderance of **system-level barriers**. This need for system-level transformation is echoed in the challenge defined by study participants: *“How might we help patients to have early, easy access to health care advice and treatment, because we want to address the disease throughout the spectrum, not just at the urgent/late stage?”* The proposed solutions are also all system-level changes.

The clear connections between insights derived in the ‘Empathize’ stage and solutions proposed during the ‘Ideate’ stage show the success of the co-design process in inspiring empathy-driven solutions. For instance, the importance of family, social, and mental health supports translated to an idea for a PAD patient support group. Lack of awareness about PAD in primary care, as noted by patients, and limited education on PAD, as noted by clinicians, translated into ideas for PAD education campaigns, improved education at the medical student level, and improved electronic consultation and referral processes that will make it easier for family physicians or other care providers to refer patients with symptoms, risk factors for PAD, or known PAD to specialty clinics.

There is evidence regarding the effectiveness of similar system-level interventions. For example, an international, prospective cohort study revealed that, amongst patients followed in specialty clinics for new or exacerbated symptoms of PAD, 89% of patients were on an antiplatelet and 83% on a statin.(30) This is compared to the general PAD population, where only approximately one-third of patients were using appropriate secondary preventative therapies.(14,31–33). Therefore, increased access to specialty services should improve outcomes.

Another example is the effectiveness of systematic smoking cessation programs, for example, the Ottawa Model for Smoking Cessation (OMSC).(34) This model standardizes processes to identify smokers, and provides smoking cessation interventions (counseling, discounted smoking cessation medications, etc.) and follow-up. It results in clinically and statistically significant differences in long-term abstinence.(34,35) Furthermore, it significantly lowers healthcare consumption (e.g. rates of all-cause hospital readmissions, smoking-related readmissions, and all-cause emergency department visits).(36)

A final example is evidence for peer support interventions for patients with various health conditions, including chronic disease such as coronary artery disease and stroke.(37–40) For example, patients with coronary artery disease who participated in peer support programs did better at sustaining physical activity and smoking cessation, and had more social support from non-family members.(37,38) Stroke survivors considered peer support valuable because it facilitated the sharing of experiences, social comparison, vicarious learning, and increased motivation.(39) Peer support is also effective among groups that are hardly reached through conventional approaches, whether that be due to individual, demographic or cultural-environmental factors.(41)

Despite the evidence in support of these interventions, they continue to be underutilized or underprioritized, especially for PAD patients. This study urges us to reevaluate these gaps. Many of the solutions proposed by workshop participants target these evidence-based, system-level interventions–through improved funding support, institutional support, outreach efforts and technological applications. Some of these solutions involve expanding or refining existing processes, while others would disrupt existing processes or need to be built from the ground up. As we proceed with this work, the prioritization index will guide us in selecting the solutions that we expect to be the most feasible and impactful. These solutions will be co-produced with our patient and public partners.

Finally, although DT methodology has been applied in the healthcare sector, we have few examples where the methods used were thorough. For example, in one systematic review of design thinking in healthcare, only 6 of 24 studies conducted contextual observations of users during the needs assessment phase (some only reported literature review, and possibly expert consultation, as their needs assessment steps); 0 studies reported a brainstorming stage; 10 studies did not use low-fidelity prototypes; and some reported a small number of iterations.(4) By being systematic and thorough in following the co-design and DT process, we also hope to advance the use of DT in health research and improvement.

### Limitations

Our study has several limitations. First, it is specific to our local context; not all centers will have access to specialty services for PAD as is available at our center, while other centers may have more system-level interventions in place (i.e., OMSC, structured exercise programs, etc.). Second, all patient participants are currently connected with specialty Vascular Medicine services. Insights may have been further enriched by also recruiting patients who are not connected with specialty services; however, our patient participants discussed their care journeys starting even before their PAD diagnoses, therefore this does capture some of that perspective. Third, the ‘Design Thinking Workshop’ was attended by primarily hospital-based professionals. The participation of more community-based professionals may have enriched our contextual understanding. Fourth, the idea prioritization survey distributed to workshop participants, especially questions regarding the feasibility of ideas, may have required a familiarity with the healthcare system beyond that of most participants. It was also only distributed in an online format, which may not have been accessible to some participants. Iterative versions of this survey will be conducted using alternative methods to inform the next stages of our work. Finally, due to the COVID-19 pandemic, the interviews and workshop were conducted virtually; we acknowledge that this has advantages and disadvantages.(42) In addition to using Google Docs, use of other online tools such as online whiteboards may have improved visualization, which in turn may have resulted in a richer depiction of the problem’s complexity and improved creativity.

## Conclusions

Underdiagnosis and undertreatment of PAD is a persistent issue. This study demonstrates how DT methodology can be applied to “wicked”–complex and ambiguous– problems, to systematically develop human-centered solutions. In exploring the lived experience of those most affected by PAD and those delivering PAD care, this study emphasizes the importance of system-level interventions to improve diagnosis and secondary prevention of PAD. The onus cannot be placed on individuals or clinicians only. In the next stages of this study, we will use the results of this co-design process to iteratively implement, evaluate and optimize some of the proposed solutions.

## Data Availability

All data produced in the present study are available upon reasonable request to the authors.

## REFERENCES

1. Jones PH. Design for care: innovating healthcare experience. Brooklyn, N.Y: Rosenfeld Media; 2013. 356 p.

2. Rittel HWJ. Dilemmas in a general theory of planning.

3. Buchanan R. Wicked Problems in Design Thinking. Des Issues. 1992;8(2):5.

4. Altman M, Huang TTK, Breland JY. Design Thinking in Health Care. Prev Chronic Dis. 2018 Sep 27;15:E117.

5. History [Internet]. IDEO | Design Thinking. [cited 2022 Jul 16]. Available from: https://designthinking.ideo.com/history

6. Ku B, Lupton E. Health design thinking: creating products and services for better health. Second edition, revised and expanded. Cambridge: The MIT Press; 2022.

7. Fowkes FGR, Rudan D, Rudan I, Aboyans V, Denenberg JO, McDermott MM, et al. Comparison of global estimates of prevalence and risk factors for peripheral artery disease in 2000 and 2010: a systematic review and analysis. The Lancet. 2013 Oct 19;382(9901):1329–40.

8. Song P, Fang Z, Wang H, Cai Y, Rahimi K, Zhu Y, et al. Global and regional prevalence, burden, and risk factors for carotid atherosclerosis: a systematic review, meta-analysis, and modelling study. Lancet Glob Health. 2020 May 1;8(5):e721–9.

9. Meru AV, Mittra S, Thyagarajan B, Chugh A. Intermittent claudication: An overview. Atherosclerosis. 2006 Aug 1;187(2):221–37.

10. Kinlay S. Management of Critical Limb Ischemia. Circ Cardiovasc Interv. 2016 Feb;9(2):e001946.

11. Torbjörnsson E, Ottosson C, Blomgren L, Boström L, Fagerdahl AM. The patient’s experience of amputation due to peripheral arterial disease. J Vasc Nurs. 2017 Jun 1;35(2):57–63.

12. Epidemiology of Peripheral Artery Disease | Circulation Research [Internet]. [cited 2022 Jul 16]. Available from: https://www-ahajournals-org.libaccess.lib.mcmaster.ca/doi/full/10.1161/CIRCRESAHA.116.303849

13. Eikelboom JW, Connolly SJ, Bosch J, Dagenais GR, Hart RG, Shestakovska O, et al. Rivaroxaban with or without Aspirin in Stable Cardiovascular Disease. N Engl J Med. 2017 Oct 5;377(14):1319–30.

14. Li B, Rizkallah P, Eisenberg N, Forbes TL, Roche-Nagle G. Rates of Intervention for Claudication versus Chronic Limb-Threatening Ischemia in Canada and United States. Ann Vasc Surg. 2022 May;82:131–43.

15. Hirsch AT, Criqui MH, Treat-Jacobson D, Regensteiner JG, Creager MA, Olin JW, et al. Peripheral Arterial Disease Detection, Awareness, and Treatment in Primary Care. JAMA. 2001 Sep 19;286(11):1317–24.

16. Bhagirath VC, Nash D, Wan D, Anand SS. Building Your Peripheral Artery Disease Toolkit: Medical Management of Peripheral Artery Disease in 2022. Can J Cardiol. 2022 May 1;38(5):634–44.

17. Vart P, Coresh J, Kwak L, Ballew SH, Heiss G, Matsushita K. Socioeconomic Status and Incidence of Hospitalization With Lower-Extremity Peripheral Artery Disease: Atherosclerosis Risk in Communities Study. J Am Heart Assoc. 6(8):e004995.

18. de Jager E, Gunnarsson R, Ho YH. Disparities in Advanced Peripheral Arterial Disease Presentation by Socioeconomic Status. World J Surg. 2022 Jun 1;46(6):1500–7.

19. Moll S, Wyndham-West M, Mulvale G, Park S, Buettgen A, Phoenix M, et al. Are you really doing ‘codesign’? Critical reflections when working with vulnerable populations. BMJ Open. 2020 Nov;10(11):e038339.

20. Roberts JP, Fisher TR, Trowbridge MJ, Bent C. A design thinking framework for healthcare management and innovation. Healthcare. 2016 Mar 1;4(1):11–4.

21. An Introduction to Design Thinking: Process Guide [Internet]. Hasso Plattner Institute of Design; 2010. Available from: https://web.stanford.edu/∼mshanks/MichaelShanks/files/509554.pdf

22. Bazzano AN, Martin J, Hicks E, Faughnan M, Murphy L. Human-centred design in global health: A scoping review of applications and contexts. PLOS ONE. 2017 Nov 1;12(11):e0186744.

23. Oliveira M, Zancul E, Fleury AL. Design thinking as an approach for innovation in healthcare: systematic review and research avenues. BMJ Innov [Internet]. 2021 Apr 1 [cited 2022 Jul 16];7(2). Available from: https://innovations-bmj-com.libaccess.lib.mcmaster.ca/content/7/2/491

24. Kumar V. 101 Design Methods: A Structured Approach for Driving Innovation in Your Organization. John Wiley & Sons; 2012. 336 p.

25. Braun V, Clarke V. Using thematic analysis in psychology. Qual Res Psychol. 2006 Jan;3(2):77–101.

26. Willig C, Rogers WS. The SAGE Handbook of Qualitative Research in Psychology. SAGE; 2017. 665 p.

27. What Is Insight? The 5 Principles of Insight Definition [Internet]. Thrive. 2016 [cited 2022 Jul 16]. Available from: https://thrivethinking.com/2016/03/28/what-is-insight-definition/

28. Design Kit: The Human-Centered Design Toolkit | ideo.com [Internet]. [cited 2022 Jul 16]. Available from: https://www.ideo.com/post/design-kit

29. Turning Insights into Opportunities [Internet]. Bridge Innovate. [cited 2022 Jul 16]. Available from: https://www.bridgeinnovate.com/blog/2017/12/19/turning-insights-into-opportunities

30. Adherence to Guideline-Recommended Therapy—Including Supervised Exercise Therapy Referral—Across Peripheral Artery Disease Specialty Clinics: Insights From the International PORTRAIT Registry [Internet]. [cited 2022 Jul 16]. Available from: https://www.ahajournals.org/doi/epdf/10.1161/JAHA.119.012541

31. Kaplovitch E, Collins A, McClure G, Tse R, Bhagirath V, Chan N, et al. Medical Therapy Following Urgent/Emergent Revascularization in Peripheral Artery Disease Patients (Canadian Acute Limb Ischemia Registry [CANALISE I]). CJC Open. 2021 Nov 1;3(11):1325–32.

32. Secondary Prevention and Mortality in Peripheral Artery Disease [Internet]. [cited 2022 Jul 16]. Available from: https://www-ahajournals-org.libaccess.lib.mcmaster.ca/doi/epdf/10.1161/CIRCULATIONAHA.110.003954

33. Berger JS, Ladapo JA. Underuse of Prevention and Lifestyle Counseling in Patients With Peripheral Artery Disease. J Am Coll Cardiol. 2017 May 9;69(18):2293–300.

34. Reid R, Mullen KA, Slovinec D’Angelo M, Aitken D, Papadakis S, Haley P, et al. Smoking cessation for hospitalized smokers: An evaluation of the “Ottawa Model.” Nicotine Tob Res Off J Soc Res Nicotine Tob. 2009 Nov 1;12:11–8.

35. Reid RD, Malcolm J, Wooding E, Geertsma A, Aitken D, Arbeau D, et al. Prospective, Cluster-Randomized Trial to Implement the Ottawa Model for Smoking Cessation in Diabetes Education Programs in Ontario, Canada. Diabetes Care. 2018 Mar 1;41(3):406–12.

36. Mullen KA, Manuel DG, Hawken SJ, Pipe AL, Coyle D, Hobler LA, et al. Effectiveness of a hospital-initiated smoking cessation programme: 2-year health and healthcare outcomes. Tob Control. 2017 May;26(3):293–9.

37. Hildingh C, Fridlund B. A 3-Year Follow-Up of Participation in Peer Support Groups after a Cardiac Event. Eur J Cardiovasc Nurs. 2004 Dec;3(4):315–20.

38. Clark AM, Munday C, McLaughlin D, Catto S, McLaren A, MacIntyre PD. Peer support to promote physical activity after completion of centre-based cardiac rehabilitation: evaluation of access and effects. Eur J Cardiovasc Nurs. 2012 Dec;11(4):388–95.

39. Clark E, MacCrosain A, Ward NS, Jones F. The key features and role of peer support within group self-management interventions for stroke? A systematic review. Disabil Rehabil. 2020 Jan 30;42(3):307–16.

40. Wan X, Chau JPC, Mou H, Liu X. Effects of peer support interventions on physical and psychosocial outcomes among stroke survivors: A systematic review and meta-analysis. Int J Nurs Stud. 2021 Sep;121:104001.

41. Sokol R, Fisher E. Peer Support for the Hardly Reached: A Systematic Review. Am J Public Health. 2016 Jul;106(7):e1–8.

42. Costantino B. The Pros and Cons of Remote Design Thinking [Internet]. [cited 2022 Sep 28]. Available from: https://www.techedgegroup.com/blog/the-pros-and-cons-of-remote-design-thinking

